# A generalizable connectome-based marker of in-scan sustained attention in neurodiverse youth

**DOI:** 10.1101/2022.07.25.22277999

**Authors:** Corey Horien, Abigail S. Greene, Xilin Shen, Diogo Fortes, Emma Brennan-Wydra, Chitra Banarjee, Rachel Foster, Veda Donthireddy, Maureen Butler, Kelly Powell, Angelina Vernetti, Francesca Mandino, David O’Connor, Evelyn M. R. Lake, James C. McPartland, Fred R. Volkmar, Marvin Chun, Katarzyna Chawarska, Monica D. Rosenberg, Dustin Scheinost, R. Todd Constable

**Author notes:** Corresponding authors: Corey Horien, Magnetic Resonance Research Center, 300 Cedar St, PO Box 208043, New Haven, CT 06520-8043, Todd Constable, Magnetic Resonance Research Center, 300 Cedar St, PO Box 208043, New Haven, CT 06520-8043.

## Abstract

Difficulty with attention is an important symptom in many conditions in psychiatry, including neurodiverse conditions such as autism. There is a need to better understand the neurobiological correlates of attention and leverage these findings for individuals in healthcare settings. Nevertheless, it remains unclear if it is possible to build robust dimensional predictive models of attention in neurodiverse populations. Here, we use five datasets to identify and validate functional connectome-based markers of attention. In dataset one, we use connectome-based predictive modelling and observe successful prediction of performance on an in-scan sustained attention task in a neurodiverse sample of youth. The predictions are not driven by confounds, such as head motion. In dataset two, we find the attention network model defined in dataset one generalizes to predict in-scan attention in a separate sample of neurotypical participants performing the same attention task. In datasets three to five, we use connectome-based identification and longitudinal scans to probe the stability of the attention network across months to years in individual participants. Our results help elucidate the brain correlates of attention in neurodiverse youth and support the further development of predictive dimensional models of other clinically-relevant phenotypes.

## Introduction

Autism spectrum disorder (hereafter ‘autism’) affects approximately 1% of children in the world (Zeidan et al. 2022) and is characterized by impairments in social communication and interaction as well as restricted and repetitive behaviors and atypical responses to sensory information (American Psychiatric Association 2013). An important symptom in autism with widespread individual differences is difficulty with attention. Between ~40-80% (Gadow et al. 2006; Lee and Ousley 2006) of individuals with autism have co-occurring attention symptoms, affecting quality of life (Masi et al. 2017). In addition, other neurodiverse individuals, like those with attention-deficit/hyperactivity-disorder (ADHD) and/or the broader autism phenotype (Ingersoll 2010) also have difficulties with attention (Gerdts and Bernier 2011; American Psychiatric Association 2013). Given the impact, there has been much recent work investigating the neurobiological correlates of attention in neurodiverse populations through the use of fMRI. Of particular interest have been functional connectivity studies, in which measures of synchrony of the blood-oxygen-level-dependent (BOLD) signal are calculated between different regions of interest (Biswal et al. 1995). Group-based functional connectivity studies—comparing those with a neurodiverse condition like autism (Di Martino et al. 2013; Keehn et al. 2013; Fitzgerald et al. 2015) or ADHD (Qiu et al. 2011; Di Martino et al. 2013; Posner et al. 2013; Hoekzema et al. 2014) to neurotypical participants—have helped advance understanding of the brain correlates of attention.

While these studies have proven useful, they have largely failed to make a clinical impact. One potential reason is a lack of prediction-based studies focusing on individuals. Studies using cross-dataset prediction—building and validating models in one sample, then testing it in a separate sample (Scheinost et al. 2019)—are rare in the neuroimaging literature, despite their potential clinical utility (Gabrieli et al. 2015). Aside from clinical applications, a prediction-based approach holds promise for avoiding statistical issues hindering generalizability (Yarkoni and Westfall 2017; Yarkoni 2020) and avoids the general lack of reliability of simple association studies (Marek et al. 2022). Finally, a prediction-based framework can offer insights into populations undergoing significant developmental changes, particularly in youth (Rosenberg et al. 2018), and aligns with the goals of interrogating symptom dimensions among diverse individuals to aid further understanding of mental disorders (Insel et al. 2010).

Based on the importance of attention and the need for prediction studies focusing on individual differences, we set out to test if it is possible to build predictive models of sustained attention phenotypes based on an in-scan attention task in a sample of youth with autism and other neurodiverse conditions, as well as neurotypical controls. There are numerous reasons it might not be possible to generate a predictive model in a youth sample comprising many patients, including difficulties with task completion and issues with scan compliance (Yerys et al. 2009). In addition, some have suggested that brain differences among those with a neurodiverse condition compared to neurotypical participants (Ross and Margolis 2019) might make it difficult to dimensionally model a phenotype using measures of functional organization. Furthermore, cognitive processes like attention are supported by complex, brain-wide correlates (Kessler et al. 2016; Rosenberg, Finn, et al. 2016). Is it possible to generate a complex, brain-wide model in a sample comprising neurotypical and neurodiverse individuals?

With these factors in mind, we address three main issues in this work. We aim to 1) determine if attention-based predictive models can be generated in a neurodiverse developmental dataset, 2) test if such a model generalizes out of sample, and 3) interrogate neuroanatomy of the network model and assess the stability in individual participants across time. Using connectome-based predictive modelling (CPM) (Finn et al. 2015; Rosenberg, Finn, et al. 2016; Shen et al. 2017; Beaty et al. 2018; Greene et al. 2018; Hsu et al. 2018; Yoo et al. 2018; Rapuano et al. 2020; Rohr et al. 2020; Boyle et al. 2022), we show that we are indeed able to predict performance on an in-scan sustained attention task in novel subjects based on functional connectivity data. The predictions are robust to factors such as in-scanner head motion, Autism Diagnostic Observation Schedule (ADOS) scores, age, sex, and intelligence quotient (IQ) scores. Crucially, we find the network model generalizes out of sample, increasing confidence in the model. In line with other dimensional work in neurodiverse populations (Lake et al. 2019; Rohr et al. 2020; Xiao et al. 2021), we observe the brain correlates identified by the model are complex and distributed across broad swaths of cortical, subcortical, and cerebellar regions. Using connectome-based identification (ID) (Finn et al. 2015; Kaufmann et al. 2017; Vanderwal et al. 2017; Waller et al. 2017; Amico and Goni 2018; Graff et al. 2022; Graff et al. 2022), we perform exploratory analyses testing the longitudinal stability of the network model in individual participants. In sum, our data suggest robust network markers of attention can be generated in neurodiverse youth and add to the growing literature suggesting the power of dimensional approaches in modelling brain-behavior relationships.

## Materials and Methods

### Description of datasets

We used five independent data sets (Table 1) in this study. The first dataset consisted of youths with autism and other neurodiverse conditions (e.g., ADHD, anxiety, broader autism phenotype, bipolar disorder) as well as typically developing children and has been described previously (hereafter ‘neurodiverse sample’) (Horien et al. 2020). Participants were scanned on a 3T Siemens Prisma system. See Supplement for exclusion criteria and imaging parameters for the neurodiverse sample. A second dataset of neurotypical adults was used as a test dataset (hereafter ‘test sample’) and is described elsewhere (Rosenberg, Finn, et al. 2016). Participants were scanned on a 3T Siemens Trio TIM system.

**Table 1.** Demographic and imaging characteristics of samples used in this study. ADHD, attention-deficit/hyperactivity disorder; BAP, broader autism phenotype; ADOS, autism diagnostic observations schedule; IQ, intelligence quotient, TR, repetition time.

| Measure | Neurodiverse sample | Utah | UM | Pitt | Test sample |
| --- | --- | --- | --- | --- | --- |
| Number of participants (males) | 70 (39) | 16 (16) | 27 (4) | 44 (21) | 25 (12) |
| Number of participants with a neurodevelopmental or psychiatric condition | 33 total<br>7 = ADHD<br>2 = anxiety disorder<br>20 = autism<br>3 = BAP<br>1 = bipolar | - | - | - | - |
| Age in years, mean<br>(standard deviation) | 11.59 (2.87) | 20.23 (8.28) | 65.3 □ (6.28) | 15.23 □ (2.83) | 22.79 (3.54) |
| Time between scans in<br>years, mean (standard<br>deviation) | - | 2.54 □ (0.29) | 0.305 □<br>(0.067) | 1.76 □ (0.41) | - |
| Scan duration in<br>minutes (volumes) | 10 min (two 5 min runs;<br>600 total volumes) | 8 min (240) | 5 min (150) | 5 min (200) | 36 min task<br>(three 12 min<br>runs; 824 total<br>volumes); 6 min<br>rest (360 total<br>volumes) |
| TR in seconds | 1 | 2 | 2 | 1.5 | 1 |
| IQ, mean (standard<br>deviation) | 107.23 (15.93) | - | - | - | - |

Three additional datasets from the Consortium for Reliability and Reproducibility (CoRR) (Zuo et al. 2014) were used to assess stability of the network model: the University of Pittsburgh School of Medicine dataset, the University of Utah dataset, and the University of McGill dataset (hereafter, ‘Pitt’, ‘Utah’, and ‘UM’, respectively). Full details of the Pitt and UM datasets can be found elsewhere (Hwang et al. 2013; Orban et al. 2015). All scans were acquired using Siemens 3-T Tim Trio scanners; all participants were neurotypical.

All datasets were collected in accordance with the institutional review board or research ethics committee at each site. Where appropriate, informed consent was obtained from the parents or guardians of participants. Written assent was obtained from children ages 13–17; verbal assent was obtained from participants under the age of 13.

### gradCPT task description

Participants in the neurodiverse sample completed the gradual onset continuous performance task (gradCPT) (Esterman et al. 2013; Rosenberg et al. 2013; Rosenberg, Finn, et al. 2016). The gradCPT task is an assessment of sustained attention and inhibition abilities that has been shown to produce a range of performance scores across neurotypical participants (Esterman et al. 2013; Rosenberg et al. 2013). In the task, participants viewed grayscale images of city and mountain scenes presented at the center of the screen. In each trial, an image transitioned from one to the next through linear pixel-by-pixel interpolation. Each transition took 1000 ms. For 1000 ms the current scene transitioned from the previous scene, and for the next 1000 ms, it transitioned to the next. Subjects were told to respond by pressing a button for city scenes and to withhold button presses for mountain scenes. City scenes occurred randomly 90% of the time. As in previous studies (Esterman et al. 2013; Rosenberg, Finn, et al. 2016), accuracy was emphasized without reference to speed, and performance was quantified using *d’* (sensitivity), the participant’s hit rate minus false alarm rate. Participant *d*’ scores were calculated for scan 1 and scan 2 individually, as well as the average across both scans. Participants in the test sample also completed gradCPT with the same parameters as above, except scene transitions took 800 ms; resting-state data were also collected in this sample.

The Pitt, Utah, and UM subjects completed only resting-state scans that were spaced apart at longer time intervals (months to years between scans; Table 1).

### Preprocessing

The preprocessing strategy for the neurodiverse sample has been described previously (Greene et al. 2018; Horien et al. 2019). Preprocessing steps were performed using BioImage Suite (Joshi et al. 2011) unless otherwise indicated, and included: skull-stripping the 3D magnetization prepared rapid gradient echo (MPRAGE) images using optiBET (Lutkenhoff et al. 2014) and performing linear and non-linear transformations to warp a 268-node functional atlas from MNI space to single subject space (Greene et al. 2018). Functional images were motion corrected using SPM8 (https://www.fil.ion.ucl.ac.uk/spm/software/spm8/). Covariates of no interest were regressed from the data, including linear, quadratic, and cubic drift, a 24-parameter model of motion (Satterthwaite et al. 2013) mean cerebrospinal fluid signal, mean white matter signal, and the global signal. Data were temporally smoothed with a zero-mean unit-variance low-pass Gaussian filter (approximate cutoff frequency of 0.12□Hz). The results of skull-stripping, non-linear, and linear registrations were inspected visually after each step.

We used previously preprocessed data for the Utah, UM, Pitt, and test samples; the preprocessing approach has been described elsewhere (Horien et al. 2019; Rosenberg, Finn, et al. 2016). (See Supplement for more about how motion was controlled in all analyses in all samples.)

### Node and network definition

We used a 268-node functional atlas (Finn et al. 2015). For each participant, the mean time-course of each region of interest (“node” in graph theory) was calculated, and the Pearson correlation coefficient was calculated between each pair of nodes to achieve a symmetric 268 × 268 matrix of correlation values representing “edges” (connections between nodes) in graph theoretic terminology. We transformed the Pearson correlation coefficients to *z*-scores via a Fisher transformation and only considered the upper triangle of the matrix, yielding 35,778 unique edges for whole-brain CPM analyses.

### Connectome-based predictive modelling

To predict gradCPT performance (*d’*) from the brain data (connectivity matrices) in the neurodiverse sample, we used CPM (Shen et al. 2017). (See Supplemental Methods and Supplemental Figure 1 for more details about CPM.)

Briefly, using 10-fold cross-validation, connectivity matrices from gradCPT and *d’* scores were divided into an independent training set including subjects from 9 folds and a testing set including the left-out fold. In the training set, linear regression was used to relate edge strength to *d’*. The edges most strongly related to *d’* were selected (using a feature selection threshold of *P* = 0.05) for both a ‘high network’ (in which increased connectivity was associated with a higher *d’* score) and a ‘low network’ (in which decreased connectivity was associated with a higher *d’* score). Mean network strength was calculated in both the high and low networks, and the difference between these network strengths was calculated (‘combined network strength’), as in previous work (Greene et al. 2018):

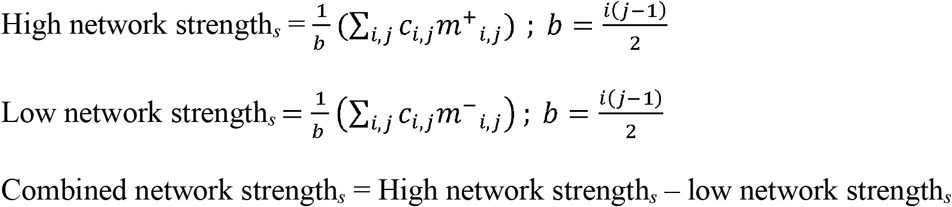

where *c* is the connectivity matrix for subject *s* and *m*^+^and *m*^−^ are binary matrices indexing the edges (*i, j*) that survived the feature selection threshold for the high or low network. (Recall that *c* and *m*^+^and *m*^−^ comprise only the upper triangle of the connectivity matrix, as specified above.) Throughout the text, we refer to the edges in the high and low network as comprising the ‘attention network.’

A linear model was then generated relating combined network strength to *d’* scores in the training data. In a final step, combined network strength was calculated for the left-out participants in the testing set, and the model was applied to generate *d*’ predictions for these left-out subjects. We conducted the main CPM analyses by constructing an average connectivity matrix per participant across the two gradCPT runs; behavioral data were averaged as well, as in previous work using gradCPT (Rosenberg, Finn, et al. 2016). (See ‘Multiverse analysis and CPM’ section below for how the effects of arbitrary choices were assessed.)

As in (Scheinost et al. 2021), model performance was assessed by comparing the similarity between predicted and observed gradCPT *d’* scores using both Spearman’s correlation (to avoid distribution assumptions) and root mean square error (defined as: 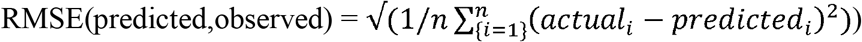. We performed 1000 iterations of a given CPM analysis and selected the median-performing model; we report this in the main text when discussing model performance. To calculate significance, we randomly shuffled participant labels and attempted to predict gradCPT *d’* scores. We repeated this 1000 times and calculated the number of times a permuted predictive accuracy was greater than the median of the unpermuted predictions to achieve a non-parametric *P*-value:

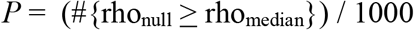

where #{rho_null_ ≥ rho_median_} indicates the number of permuted predictions numerically greater than or equal to the median of the unpermuted predictions (Scheinost et al. 2021).

### Multiverse analysis and CPM

To determine if CPM findings were robust, we used a multiverse approach to explore how results were affected by different analytical choices (Steegen et al. 2016). The goal of this approach is not to determine what CPM pipeline gives the ‘best’ prediction; rather, it is to gather converging evidence across a range of analytical scenarios to determine how modelling choices affect results. Specifically, we altered the feature selection threshold used to select significant edges (*P* = 0.05, *P* = 0.01, *P* = 0.005, *P* = 0.001); we tested CPM using a combined network model versus testing the high and low networks separately; we tested the effect of partialling out participant age, sex, IQ, Autism Diagnostic Observation Schedule (ADOS) (Lord et al. 2012) calibrated severity score, and head motion in the feature selection step (as described previously) (Scheinost et al. 2021; Dufford et al. 2022); and we also built models using data from gradCPT run 1, gradCPT run 2, and average gradCPT data. To ensure this approach did not result in false positives, the Benjamini–Hochberg procedure (Benjamini and Hochberg 1995) was applied to control for multiple comparisons (correcting for 12 tests).

### Testing generalizability of the attention network

To assess if the network model of attention generalized out of sample, we defined a consensus high attention network and consensus low attention network as edges that appear in at least 6/10 folds in 600/1000 iterations of CPM. This resulted in 922 edges in the high attention network and 896 edges in the low attention network. Using the combined network strength in the consensus networks (as above for CPM), we determined model coefficients across the neurodiverse sample, as in (Rosenberg, Finn, et al. 2016; Ju et al. 2020; Dufford et al. 2022). We then applied the network masks and model coefficients to the test sample to generate *d*’ predictions.

As above for CPM analyses, model performance was assessed by comparing the similarity between predicted and observed gradCPT *d’* scores using both Spearman’s correlation and by calculating RMSE. Non-parametric *P*-values were computed as for CPM. As above, we used a similar multiverse approach to ensure results were not being driven by subject age, sex, or head motion; the Benjamini–Hochberg (1995) procedure was again used to control for multiple comparisons. To further ensure results were robust, we tested a range of summary networks of varying sizes (i.e., from stringent cases where an edge must appear in 10/10 folds and 1000/1000 iterations, to more liberal thresholds where an edge must appear in 3/10 folds and 300/1000 iterations).

### Connectome-based ID

To test the stability of the predictive model over time in a given individual, we used connectome-based ID (Finn et al. 2015) and the Pitt, Utah, and UM samples. (See Supplemental Figure 2 for a schematic and more about connectome-based ID.) Briefly, after selecting only the edges in the high and low networks (i.e., the same consensus edges used in the cross-dataset test—the 922 and 896 edges in the high and low attention networks, respectively), a database was created consisting of all subjects’ matrices from scan 1. In an iterative process, a connectivity matrix from a given subject was then selected from scan 2 and denoted as the target. Pearson correlation coefficients were calculated between the target connectivity matrix and all the matrices in the database. If the highest Pearson correlation coefficient was between the target subject in one session and the same subject in the second session (i.e. within-subject correlation > all other between subject correlations), this was recorded as a correct identification. The process was repeated until identifications had been performed for all subjects and database-target combinations. We averaged both database-target pairs (because these can be reversed) for a dataset to achieve an average ID rate. To calculate *P*-values, we randomly shuffled subject identities and reperformed ID for 1000 iterations and compared the actual ID rates to this null distribution (Finn et al. 2015; Horien et al. 2018; Horien et al. 2019):

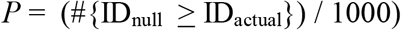

where #{ID_null_ ≥ ID_actual_} indicates the number of permuted ID rates numerically greater than or equal to the actual ID rate obtained using the original data.

We also assessed if connections inside the attention network were more or less stable than connections in the rest of the brain. We generated 1000 summary networks comprising edges outside of the consensus attention network (and the same size as the high and low attention networks). Connectome-based ID was performed using the random networks, and we compared ID results to those obtained using the original consensus attention network. *P*-values were obtained as follows:

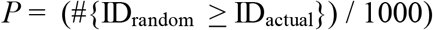

where #{ID_random_ ≥ ID_actual_} indicates the number of random ID rates numerically greater than or equal to the actual ID rate obtained using the original data.

In the Pitt dataset, incomplete scan coverage during the functional runs resulted in 158/922 and 145/896 edges missing in the high and low networks, respectively; we performed ID with the remaining edges.

### Code and data availability

Preprocessing was carried out using software freely available here: (https://medicine.yale.edu/bioimaging/suite/). CPM code is available here: (https://github.com/YaleMRRC/CPM). The parcellation, the attention network models, and the connectome-based ID code are available here: (https://www.nitrc.org/frs/?group_id=51). Data from the longitudinal samples are openly available through CoRR (http://fcon_1000.projects.nitrc.org/indi/CoRR/html/).

## Results

### Prediction of in-scan attention scores in the neurodiverse sample

In the neurodiverse sample, there were no differences between neurodiverse and neurotypical participants in in-scanner head motion (t(68) = 0.77, *P* = 0.4437) or gradCPT *d’* (t(68) = −0.60, *P* = 0.5487). Across the sample, we observed that motion and *d’* were negatively correlated (r = −0.35, *P* = 0.0034); we hence adopted a rigorous motion control strategy for the duration of the analyses to ensure motion was not driving the brain-behavior models (Supplemental Methods).

Next, using CPM, we built a model using within-dataset cross-validation to predict unseen participants’ gradCPT *d’* scores from functional connectivity data. The model successfully predicted gradCPT *d*’ scores in the neurodiverse dataset (Spearman’s rho = 0.54, RMSE = 0.78, *P* = 0.0001; Figure 1A).

**Figure 1.**
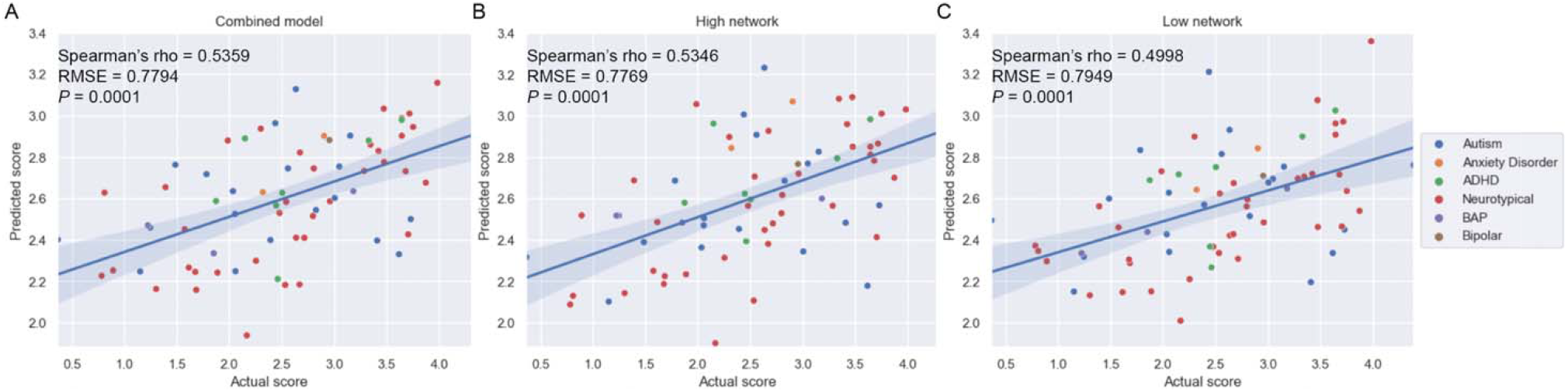
In-scan sustained attention task performance (gradCPT *d’*) can be predicted in a neurodiverse sample using CPM. A) Results from the combined network model. B) Results from the high network. C) Results from the low network. For all plots, actual gradCPT *d*’ scores are indicated on the x-axis; predicted scores, on the y-axis. A regression line and 95% confidence interval are shown. ADHD, attention-deficit/hyperactivity disorder; BAP, broader autism phenotype; *P* = *P*-value; RMSE, root mean square error.

To assess the robustness of the *d*’ prediction, we used a multiverse approach to explore how results were affected by different analytical choices (Steegen et al. 2016). We stress the point of this approach is not to determine what pipeline gives the ‘best’ prediction performance; it is instead to gather converging evidence across a range of analytical scenarios to determine the extent to which arbitrary choices affect CPM results.

As the choice of feature selection threshold is arbitrary, we started by testing a range of thresholds, while still controlling for motion. We were able to significantly predict *d*’ scores in all cases (feature selection threshold of 0.01: Spearman’s rho = 0.50, RMSE = 0.79, *P* = 0.0001; feature selection threshold of 0.005: Spearman’s rho = 0.45, RMSE = 0.80, *P* = 0.002; feature selection threshold of 0.001: Spearman’s rho = 0.43, RMSE = 0.81, *P* = 0.0001). Interestingly, all models performed quite well, but when more stringent feature selection thresholds were applied (i.e., fewer edges were included in a model), performance decreased.

We next performed analyses adjusting for participant age, sex, IQ, and ADOS score. Each pipeline demonstrated similar prediction performance of *d*’ (age-adjusted model: Spearman’s rho = 0.47, RMSE = 0.80, *P* = 0.001; sex-adjusted model: Spearman’s rho = 0.52, RMSE = 0.77, *P* = 0.0001; IQ-adjusted model: Spearman’s rho = 0.49, RMSE = 0.96, *P* = 0.003; ADOS-adjusted model: Spearman’s rho = 0.53, RMSE = 0.77, *P* = 0.0001). Models were also built for gradCPT scan 1 and gradCPT scan 2 separately (scan 1: Spearman’s rho = 0.31, RMSE = 0.85, *P* = 0.031; scan 2: Spearman’s rho = 0.27, RMSE = 0.87, *P* = 0.073). Prediction performance dropped in this case, echoing recent results that more data fed into predictive models results in higher accuracies (Taxali et al. 2021).

Finally, the choice to model attention using a ‘combined network’ (used in all analyses above) is also arbitrary. We repeated the CPM prediction of *d*’ scores and tested the high and low networks. We again observed similar *d*’ prediction in both cases (high network: Spearman’s rho = 0.53, RMSE = 0.78, *P* = 0.0001; low network: Spearman’s rho = 0.50, RMSE = 0.79, *P* = 0.0001; Figure 1B-C).

In all, these results suggest attention prediction is robust in this sample and is not driven by potential confounding factors.

### External validation of the attention network

Overfitting—deriving statistical patterns specific to noise in a sample—is a constant concern in machine learning studies. The ultimate test is to assess how well a model works in an independent dataset; we perform such a test here. We determined which edges tended to contribute consistently to successful prediction (922 in the high network and 896 in the low network, 1818 edges total) and built a consensus model in the neurodiverse sample (Methods). Applying the model to the test sample comprising subjects completing the same gradCPT task, we observed successful prediction of *d*’ scores (Spearman’s rho = 0.65, RMSE = 1.059 *P* = 0.0006; Figure 2A).

**Figure 2.**
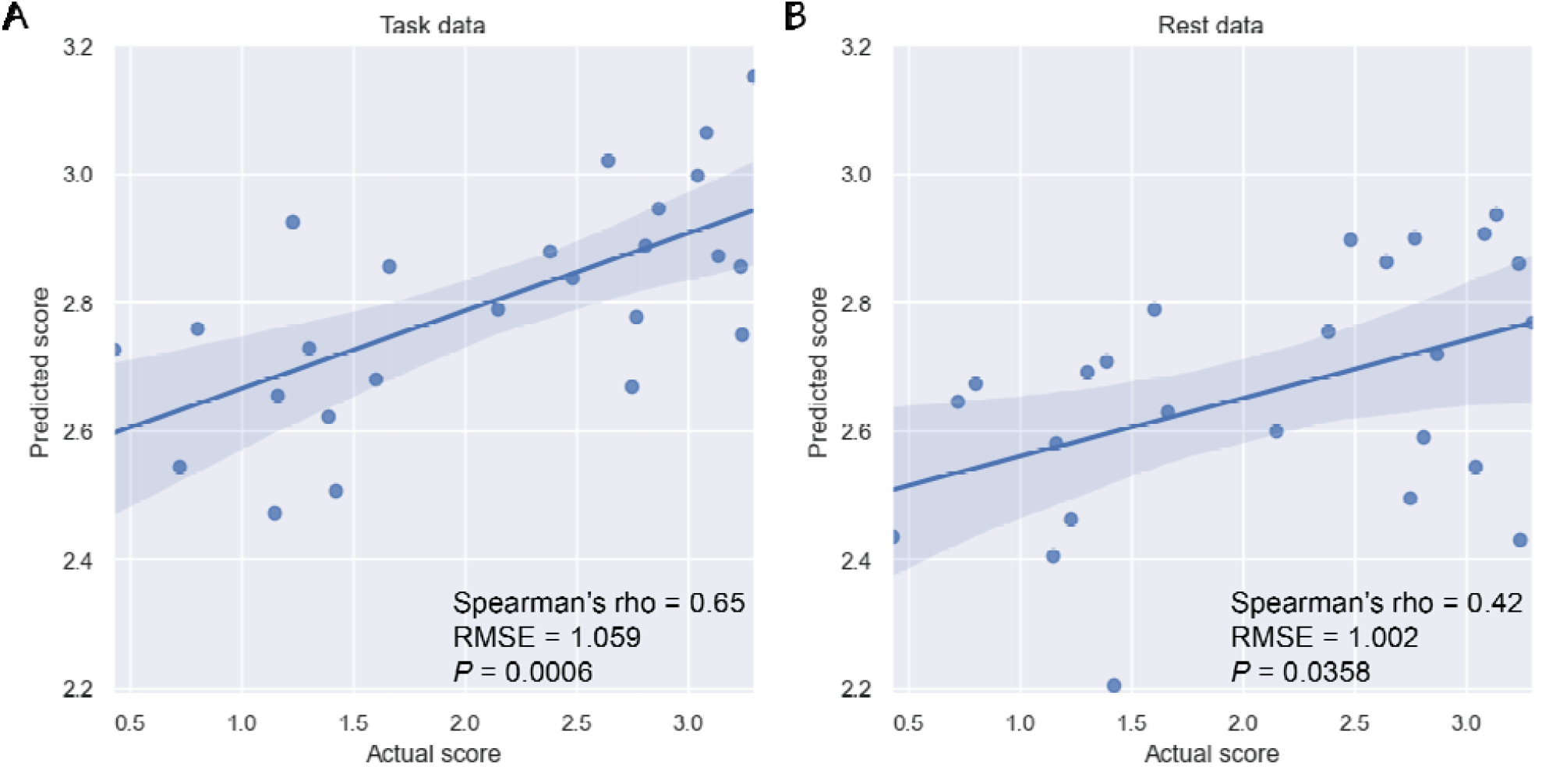
Generalization of the attention network to an independent sample. A) Results using task data. B) Results using rest data. For all plots, actual gradCPT *d*’ scores are indicated on the x-axis; predicted scores, on the y-axis. A regression line and 95% confidence interval are shown. Abbreviations: RMSE, root mean square error; *P* = *P*-value.

We repeated analyses controlling for several other variables, adjusting for in-scanner head motion (Spearman’s rho = 0.67, *P* = 0.0008), participant sex (Spearman’s rho = 0.61, *P* = 0.0014), as well as age (Spearman’s rho = 0.65, *P* = 0.0009), and observed similar results. In addition, when we tested both the high and low network separately, we found each of these networks predicted *d*’ scores (high network: Spearman’s rho = 0.59, RMSE = 1.094, *P* = 0.021; low network: Spearman’s rho = 0.59, RMSE = 1.03, *P* = 0.0025). In addition, we tested whether the attention network could be used to predict *d*’ scores from resting-state data in the adult sample. We again found the model generalized (Spearman’s rho = 0.42, RMSE = 1.002, *P* = 0.0358; Figure 2B).

We further tested the stability of results by altering how consistently an edge had to appear across CPM iterations to be included in the summary attention network (Methods). This resulted in 8 attention summary networks, ranging from ~100-3000 edges. In 7/8 cases, the attention network generalized to predict *d*’ scores (range of Spearman’s rho = 0.50-0.66; all *P* < 0.0104 after FDR correction; Supplemental Table 1). The only summary network that did not predict *d*’ score (Spearman’s rho = 0.23, *P* = 0.27) was quite small (~100 edges), approximately an order of magnitude smaller than the original attention network tested above (and other networks that have generalized) (Rosenberg, Finn, et al. 2016; Greene et al. 2018; Yip et al. 2019). These results suggest generalization in this sample is robust to the arbitrary choices made when defining a summary model.

### Neuroanatomy of CPM predictive networks

We next performed post-hoc visualizations to localize brain connections contributing to the model. Together, the 1818 total edges comprise 5.08% of the connectome. Similar to other CPM models (Rosenberg, Finn, et al. 2016; Beaty et al. 2018; Greene et al. 2018; Lake et al. 2019; Ju et al. 2020; Dufford et al. 2022), the predictive edges in the high and low networks comprise complex, distributed networks spanning the entire brain (Figure 3A-B). In line with task demands, additional visualizations at the network level (Figure 3C-D) revealed connections involving subcortical, cerebellar, and visual networks were particularly important. In the high attention network, for example, the top three network pairs containing the greatest proportion of edges involved subcortical, cerebellar, and visual networks. In the low attention network, a network pair involving the cerebellum (the cerebellar-frontoparietal network) contained the greatest proportion of edges. For completeness, we present the matrices in Figure 3C-D in Supplemental Figure 3 without normalizing by network size; subcortical, visual, and cerebellar networks again tended to harbor large numbers of edges. (See also Supplemental Figure 4 for an additional visualization using circle plots and glass brains.)

**Figure 3.**
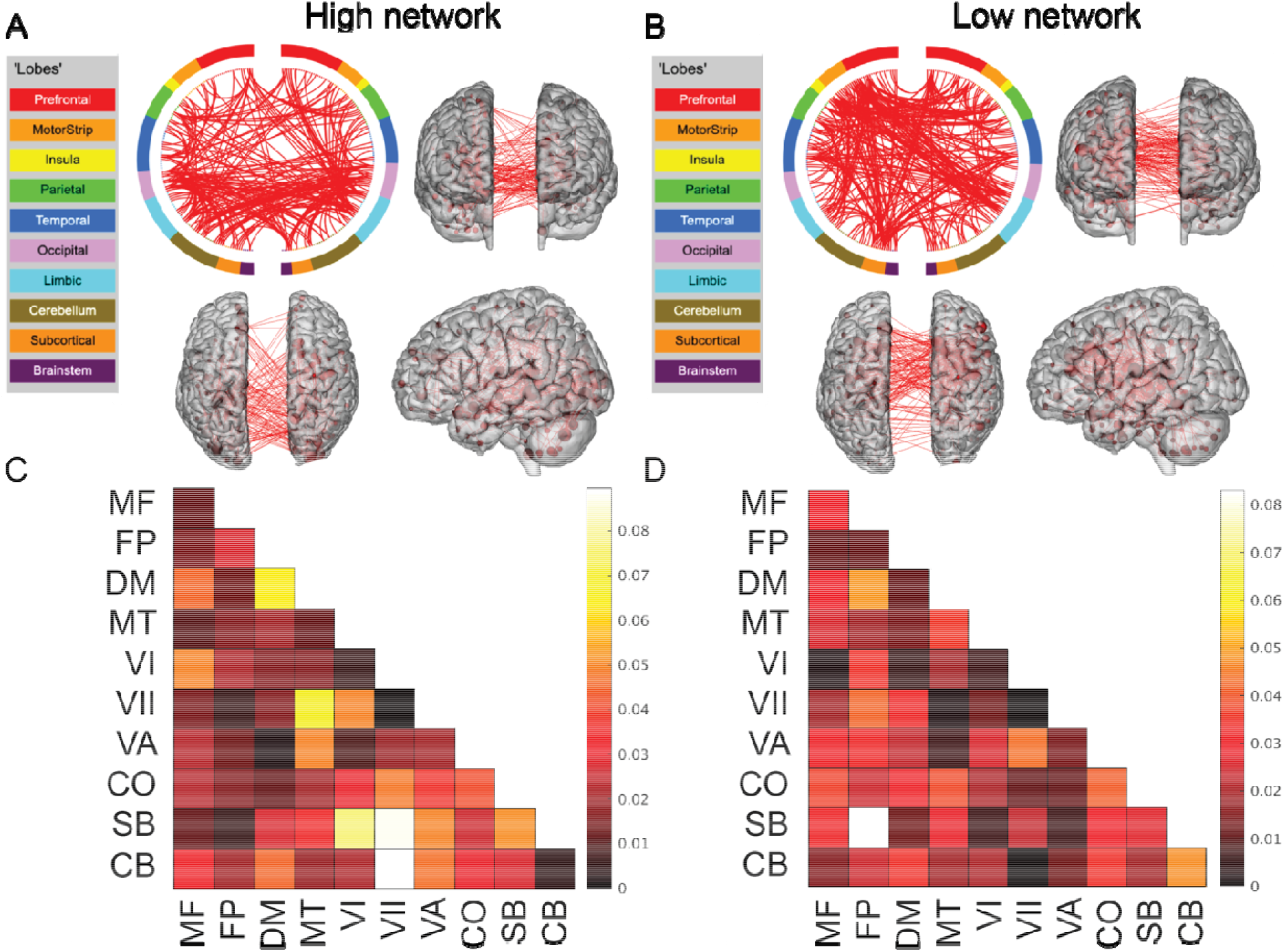
Neuroanatomy of CPM predictive networks. A) The consensus high network. B) The consensus low network. For both A) and B): a circle plot is shown in the upper left. The top of the circle represents anterior; the bottom, posterior. The left half of the circle plot corresponds to the left hemisphere of the brain. A legend indicating the approximate anatomic ‘lobe’ is shown to the left. The same edges are plotted in the glass brains as lines connecting different nodes (red circles); in these visualizations, nodes are sized according to degree, the number of edges connected to that node. Note that to aid in visualization, we have thresholded the matrices to only show nodes with a degree threshold > 15 (unthresholded circle plots and glass brains are shown in Supplemental Figure 4). C) Matrix of the consensus high network. D) Matrix of the consensus low network. For both C) and D): the proportion of edges in a given network pair; data have been corrected for differing network size. MF, medial frontal; FP, frontoparietal; DM, default mode; MT, motor; VI, visual I; VII, visual II; VA, visual association; CO, cingulo-opercular; SB, subcortical; CB, cerebellum.

### Individual-level stability of predictive network model

An ultimate goal of using individual-level approaches in the clinic is to infer future outcomes based on current data. Because the attention network was defined in relation to a state-based cognitive process that itself fluctuates (Cohen and Maunsell 2011; Esterman et al. 2013; Esterman et al. 2014; Rosenberg et al. 2015; Terashima et al. 2021), it is possible individual connectivity patterns in the attention network might change over time. Hence, we conducted an exploratory analysis using connectome-based ID and longitudinal datasets with months to years between scans, asking: are connections in the attention network stable enough within individuals to identify a participant from a group?

Across the three longitudinal samples, we observed that the attention network results in ID rates well-above chance (ID rate range: 53.4 - 93.5%; *P* < 0.0001 across all samples; Figure 4A-B). Specifically, ID rates were high when there were months between scans (UM dataset; 92.6% and 81.5% in the high and low attention networks, respectively) and when there were years between scans (as for Utah and Pitt datasets; Utah high attention network = 84.4%, Utah low attention network = 81.3%; Pitt high attention network = 53.4%, Pitt low attention network = 62.5%). These results suggest participants retain their unique connectivity signatures in the attention network.

**Figure 4.**
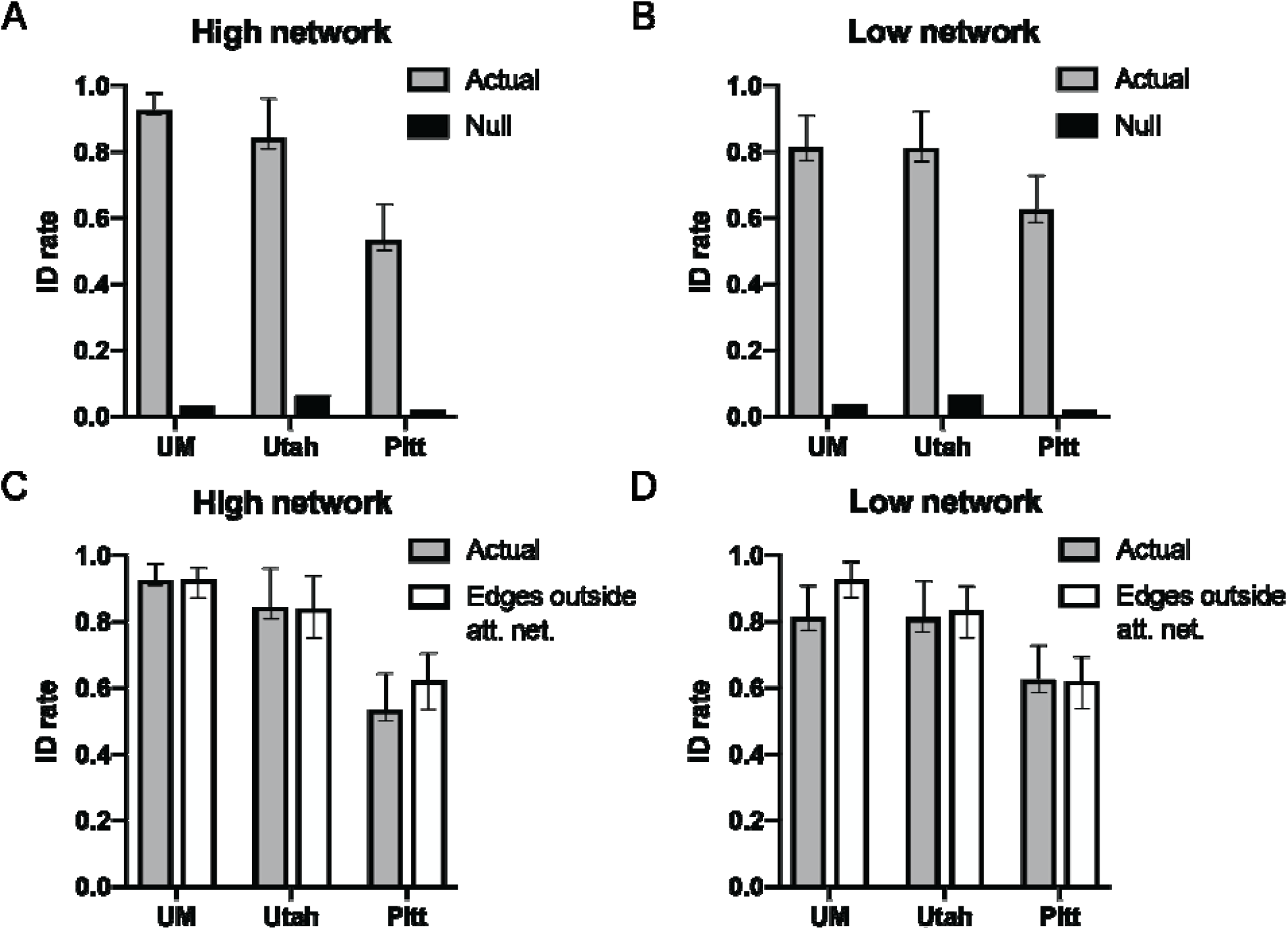
The attention network is stable at the individual level across months and years. For panels A) and B) results are shown for the high and low networks, respectively, comparing ID rates obtained using edges in the attention network (‘Actual’) compared to chance levels when participant labels have been permuted (‘Null’). In A) and B), the rightmost bar is the mean ID rate from 1000 iterations of permutation testing. For panels C) and D) results are shown for the high and low networks, respectively, comparing ID rates obtained using edges in the attention network (‘Actual’) compared to ID rates obtained when using edges not included in the attention network (‘Edges outside att. net.’). For A-D): each dataset is indicated below the x-axis; the y-axis is the ID rate, presented as a fraction from 0 (no subjects correctly identified) to 1 (all subjects correctly identified). Error bars correspond to 95% confidence intervals obtained via bootstrapping. Note the actual ID rates shown in plots A) and B) (the grey bars) are reproduced in plots C) and D). Att. net., attention network; ID rate, identification rate; Pitt, University of Pittsburgh sample; UM, University of Montreal sample.

We next examined the relationship between in-scanner head motion and within-participant correlations in connectivity strength in the attention network. Across all three samples, we found there were no statistically significant relationships between within-participant correlation scores and head motion in 5/6 cases (range of rho values: −0.096 - 0.123; *P* > 0.53 across all samples; Supplemental Table 2). The only statistically significant relationship we observed was in Pitt, and there was a negative association (high attention network: rho = −0.36, *P* = 0.0153), indicating higher head motion in this sample was associated with lower within-subject self-correlation scores (in line with previous results) (Horien et al. 2018; Graff et al. 2022). These findings suggest the high ID rates are not being driven by head motion.

Lastly, we used connectome-based ID to determine how the attention network compared to connections in the rest of the brain. Generating 1000 summary networks comprising edges outside of the consensus attention network (and of the same size), we observed no differences in ID rates of the random networks compared to the attention network (ID rate range: 62 - 92.8%; all *P* > 0.49 across all samples; Figure 4C-D; see Supplemental Table 3 for exact ID rates and *P*-values for all samples). That is, the attention network itself tends to retain participant-specific connectivity signatures across months to years because the brain as a whole retains participant-specific connectivity signatures across months to years.

## Discussion

In this work, we set out to test if it was possible to generate connectome-based predictive models of attention in a neurodiverse sample of youth. Using CPM, we were able to build a predictive network model of in-scan sustained attention scores. Crucially, we found the network generalized out of sample, further suggesting the brain-behavior model we originally identified is a robust marker of attention. The network model was spatially complex, comprising connections across the brain. Lastly, we conducted exploratory analyses in three open-source samples using the network and connectome-based ID.

### The power and potential of dimensional models

Our work adds to the growing literature suggesting it is feasible to use a dimensional approach to model individual differences in brain-behavior relationships in neurodiverse youth. In addition to autism symptoms (Lake et al. 2019), groups have developed dimensional models predictive of behavioral inhibition (Rohr et al. 2020), social affect (Xiao et al. 2021), and adaptive functioning (Plitt et al. 2015) in neurodiverse samples. In all cases, the predictive models comprise complex networks, with connections spanning the entire brain (reviewed in) (Horien et al. 2022). Nevertheless, subcortical and cerebellar networks tend to emerge as major contributors in these models, regions we also observed as important in our attention network. Furthermore, similar brain areas have also been noted to play a role in attention (Rosenberg, Finn, et al. 2016; Green et al. 2017; Yoo et al. 2022), consistent with the growing recognition that subcortical and cerebellar circuits are important in mediating cognitive processes (Buckner 2013; Clark et al. 2021).

Beyond helping to hone in on brain areas involved in sustaining attention, the network identified is intriguing from a clinical standpoint. For example, it has been shown that a network connectivity model of attention (Rosenberg, Finn, et al. 2016) is sensitive to methylphenidate (Rosenberg, Zhang, et al. 2016). It is therefore possible the network identified here may help in tracking changes after administration of a therapeutic. Though more work is needed, it is generally encouraging that markers identified through dimensional analyses appear to be sensitive to clinically useful drugs. There is a need for objective, biological markers in psychiatry, and dimensional approaches could offer a framework to identify quantitative markers to help individuals clinically (McPartland 2021).

### Generalizability of the attention network and open science

Much has been written about the reproducibility crisis in biomedical science (Pashler and Wagenmakers 2012; Open Science Collaboration 2015; Baker 2016), as well as the fear results might suffer from a lack of generalizability (Yarkoni 2020), particularly in psychiatry and psychology. We emphasize the present study uses predictive modeling, which is an additional step beyond association studies, that helps reproducibility (Rosenberg, Finn, et al. 2016; Yarkoni and Westfall 2017; Rosenberg et al. 2018; Scheinost et al. 2019; Poldrack et al. 2020). Testing to ensure results are robust across samples and contexts is also imperative. This effort is especially important in neuroimaging, where hopes have been high for clinical impact, yet there has been little progress translating papers into practice (Chekroud 2017; Chekroud and Koutsouleris 2018). Further, even when findings might be clinically useful, many roadblocks stand in the way of successful implementation (Chekroud and Koutsouleris 2018). It is incumbent on researchers to test findings in multiple samples to avoid having other investigators waste time and resources.

Hence, the fact the attention network generalizes out of sample increases confidence in the original model and opens new opportunities for analysis. That is, the attention network generalized from a young, neurodiverse sample to an older, neurotypical sample. This finding is in line with the dimensional view of brain-behavior relationships (Insel et al. 2010) and also supports the notion that despite developmental changes in brain function, a ‘core’ brain network architecture associated with sustaining attention is likely present (Yoo et al. 2022). An important next step to further assess generalizability will be testing if the network can generalize to predict different aspects of attention (or other phenotypes entirely) and in different populations. To this end, we openly share the attention network model and encourage other investigators to test it widely. By sharing materials, particularly those from neurodiverse participants, we as a community can ensure our findings are relevant for all individuals.

### Individual stability of the attention network

Using connectome-based ID, it has been shown the connectome tends to be individually stable across short time scales (minutes) (Miranda-Dominguez et al. 2014; Finn et al. 2015; Kaufmann et al. 2017; Vanderwal et al. 2017; Waller et al. 2017; Amico and Goni 2018) and longer time scales (months to years) (Miranda-Dominguez et al. 2018; Horien et al. 2019; Demeter et al. 2020; Jalbrzikowski et al. 2020; Ousdal et al. 2020; Graff et al. 2022; Graff et al. 2022). Further, individual resting-state networks have tended to exhibit high stability (Finn et al. 2015). Here, we tested if a state-based, behaviorally defined network comprising edges across the brain exhibited the same degree of stability.

Encouragingly, we observed the attention network is a stable multivariate marker of connectivity over long time scales, and this did not differ from the rest of the brain in terms of stability. We interpret this finding as a positive for the field, as it suggests that networks for other phenotypes, with different neurobiological correlates, will likely exhibit a high degree of individual stability as well. Indeed, this seems to be the case, as recent work has demonstrated that predictive network models tend to have substantially higher reliability than individual functional connections (Taxali et al. 2021). Future studies could more rigorously assess how stability of the attention network relates to CPM model performance in longitudinal samples, as the datasets used here did not contain assessments of attention. Continuing to focus on the stability and reliability of our measurements is crucial (Noble et al. 2019), as a lack of reliability can continue to impede the clinical utility of fMRI (Milham et al. 2021)

### Limitations and future considerations

Compared to other open source datasets (e.g., ABCD (Casey et al. 2018), the Human Connectome Project (Van Essen et al. 2013), UK Biobank (Miller et al. 2016)), the samples used here are small. Another limitation is participants in the neurodiverse sample had fairly high IQ scores compared to the population at large (Wingate et al. 2014). While IQ was not shown to be a confounding factor in the predictive model, individuals with lower IQs may have difficulties completing the gradCPT task. Future studies could address generalizability of the task and/or network in more varied individuals. In addition, attention is a broad construct, and in this work, we focused on the ability to sustain attention. More research could be conducted to determine if it is possible to build dimensional models of other aspects of attention in neurodiverse individuals.

The longitudinal samples we used to measure stability of the model did not contain behavioral/clinical data. It is unclear if the predictive network is able to predict attention phenotypes across longer time scales. Studies with the same participants completing the gradCPT task at multiple time points could help to answer this question. Finally, we have focused on a single phenotype here. An important next step will be to use a multimodal, multidimensional framework in large numbers of individuals—incorporating numerous phenotypes and data types—to generate findings across multiple spatial and temporal scales (Lombardo et al. 2019). Such an approach holds the promise of illuminating the complex biology underlying heterogenous neurodiverse conditions.

## Conclusion

In sum, we have shown that it is possible to generate task-based predictive models of in-scan attention in a neurodiverse sample and that such a model generalizes. Results support the further development of predictive dimensional models of cognitive phenotypes and suggest that such an approach can yield stable imaging markers.

## Data Availability

All data produced in the present study are available upon reasonable request to the authors

## Acknowledgements and Disclosures

The authors thank Hedwig Sarofin and Cheryl McMurray for technical assistance during the MRI scans and Jitendra Bhawnani for technical assistance with task hardware and software. This work was supported by the National Institutes of Health (P50MH115716, T32GM007205 to CH and ASG, and TR001864 to ASG). JCM consults with Customer Value Partners, Bridgebio, Determined Health, and BlackThorn Therapeutics, has received research funding from Janssen Research and Development, serves on the Scientific Advisory Boards of Pastorus and Modern Clinics, and receives royalties from Guilford Press, Lambert, and Springer.

