## Supplemental Materials for "A generalizable connectome-based marker of in-scan sustained attention in neurodiverse youth"

**Supplemental Methods**

Exclusion criteria

The neurodiverse sample was derived from an ongoing study (Horien et al. 2020). Participants were screened over the phone for basic developmental history and MRI safety factors. Those with a history of prematurity, known genetic abnormalities, or an IQ below 70 were excluded. Diagnostic classification of autism was based on clinical best estimate diagnosis by a team of clinical psychologists and any available reports of developmental and medical history. Ninety-two participants were scanned. We required all study participants to pass visual QC after preprocessing (i.e. all skull-stripped data, linear registrations, and non-linear registrations were manually inspected), as well as to have both gradCPT runs. We excluded participants with a lack of full brain coverage during functional scans (n = 5) and participants who were determined to have failed the preprocessing pipeline during visual inspection and/or to have an imaging artifact (n = 8). Participants were also excluded due to a technical issue with the task laptop preventing proper recording of button presses (n = 7) and a participant (n = 1) was excluded due to having only one gradCPT run, along with a participant (n =1) who pressed a combination of buttons during the task preventing proper data analysis. This left 70 participants who passed visual QC and had both gradCPT runs. In addition, one participant from this pool of 70 did not have an IQ assessment; when examining the effect of IQ on CPM results, we dropped this participant from analyses. Similarly, one participant from the test sample did not have age data; we dropped this individual from analyses when examining the impact of age.

Neurodiverse sample image acquisition parameters

All subjects from the neurodiverse sample were scanned on a 3 T Siemens Prisma system at the Yale Magnetic Resonance Research center. We acquired a high-resolution T1-weighted 3D anatomical scan using a magnetization prepared rapid gradient echo (MPRAGE) sequence with the following image parameters: 208 contiguous slices acquired in the sagittal plane, repetition time (TR) = 2400 ms, echo time (TE) = 1.22 ms, flip angle = 8°, slice thickness = 1 mm, in-plane resolution = 1 mm × 1 mm, matrix size = 256 × 256. A T1-weighted 2D anatomical scan was acquired using a fast low angle shot (FLASH) sequence with the following image parameters: 75 contiguous slices acquired in the axial-oblique plane parallel to AC-PC line, TR = 440 ms, TE = 2.61 ms, flip angle = 70°, slice thickness = 2 mm, in-plane resolution = 0.9 mm × 0.9 mm, matrix size = 256 × 256.

Functional images were acquired using a multiband gradient echo-planar imaging (EPI) pulse sequence with the following image parameters: 75 contiguous slices acquired in the axial-oblique plane parallel to AC-PC line, TR = 1000 ms, TE 30 ms, voxel size = 2.0 mm^3^, flip angle = 55 degrees, slice thickness = 2 mm, bandwidth = 1894 Hz/pixel, matrix size = 110 x 110, field of view = 220 mm, multiband factor = 5.

Motion control considerations

In-scanner head motion has been shown to affect estimates of functional connectivity (Satterthwaite et al. 2012; Power et al. 2015) and brain-behavior relationships (Siegel et al. 2017). We therefore adopted a rigorous motion control strategy during scanning acquisition and in all analyses.

Specifically, all participants underwent an intensive mock scan protocol five days prior to scanning (along with a refresher training period on the day of the scan). We have previously shown in this same sample that the mock scan protocol significantly lowers motion artifact (Horien et al., 2020). In the present paper, such an approach led to 95.7% of the sample (67/70 participants) containing data with a mean frame-to-frame displacement (FFD) < 0.2 mm, a typical threshold used for determining high versus low-motion data in youth and/or those with a mental health condition (Yip et al. 2019; Ju et al. 2020; Lichenstein et al. 2021).

In addition, similar to other recent CPM papers (i.e., Lake et al., 2019; Scheinost et al., 2021; Dufford et al., 2022), steps were taken during CPM and connectome-based ID analyses to limit the effects of motion. Specifically, we adjusted the CPM model for each participant’s mean FFD over the course of gradCPT and found that motion was not driving predictions (i.e., the model still successfully predicted gradCPT *d*’ scores when controlling for motion (Spearman’s rho = 0.54, RMSE = 0.78, *P* = 0.0001). Next, when testing if the network model generalized in the test sample, we again controlled for in-scanner head motion and found that models were not confounded by mean FFD (i.e., successful prediction was again achieved, Spearman’s rho = 0.67, *P* = 0.0008). In addition, the fact that the model from the neurodiverse sample generalizes to predict *d*’ in the test sample (with slightly faster trials; neurodiverse inter-trial interval of 1000 ms, test sample inter-trial interval of 800 ms) increases confidence that model success is not due to participant head motion yoked to stimuli presentation.

In connectome-based ID, we focused our analyses on only the low-motion longitudinal subjects previously described in Horien et al. (2019). These participants have previously been used in whole-brain and canonical network-based ID analyses, and it was shown that connectome-based ID results were not driven by head motion. From this low-motion sample (i.e., all participants had a mean FFD < 0.1 mm for all resting-state scans) we further considered how within-participant self-correlations derived from the connectome-based ID process related to in-scanner head motion in the present analyses. Across all three datasets, we found there were no statistically significant relationships between within-participant correlation scores and head motion in 5/6 cases (range of rho values: -0.0964 - 0.1236; *P* > 0.53 across all samples; Supplemental Table 2). The only statistically significant relationship we observed was in Pitt, and there was a negative association (high attention network: rho = -0.3636, *P* = 0.0153), indicating higher head motion in this sample was associated with lower within-subject self-correlation scores indicating higher head motion in this sample was associated with lower within-subject self-correlation scores (in line with previous results) (Horien *et al.* 2018; Graff *et al.,* 2022). These results suggest that head motion is not acting as a confound in the connectome-based ID results.

In sum, while head motion is always a concern in functional connectivity analyses of brain-behavior relationships, the present data suggest it is not driving the findings described here.

Connectome-based predictive modelling (CPM)

To predict gradCPT d’ scores from connectivity matrices, we used CPM (e.g. (Shen et al. 2017) (Supplemental Figure 1). In CPM, a set of connectivity matrices and behavioral variables are considered (one each per participant; first box). Data are divided into a training and testing set for cross-validation. In the training set, linear regression is used to relate edge strength to *d’* (second box). The edges most strongly related to *d’* are selected (third box) for both a ‘high network’ (in which increased connectivity is associated with a higher *d’* score) and a ‘low network’ (in which decreased connectivity is associated with a higher *d’* score). Mean network strength is calculated in both the high and low networks (fourth box, note the original work of Shen et al. (2017) had users sum the selected edges, and this is indicated on the title of the plot; we calculate mean network strength in the present work) and the difference between these network strengths is calculated (‘combined network strength’), as in previous work (Greene et al. 2018):

High network strength*_s_* = $\frac{1}{b}(\sum_{i,j} c_{i,j}{m^{+}}_{i,j}) ;b=\frac{i\left( j-1 \right)}{2}$

Low network strength*_s_* $=\frac{1}{b}\left( \sum_{i,j} c_{i,j}{m^{-}}_{i,j} \right) ;b=\frac{i\left( j-1 \right)}{2}$

Combined network strength*_s_* = High network strength*_s_* – low network strength*_s_*

where $c$ is the connectivity matrix for subject *s*, $m^{+}$and $m^{-}$are binary matrices indexing the edges ($i,j)$ that survived the feature selection threshold for the high or low network. (Recall that $c$ and $m^{+}$and $m^{-}$ comprise only the upper triangle of the connectivity matrix, as specified in the main text.)

A linear model is then generated relating combined network strength to *d’* scores in the training data (fifth box). In a final step, combined network strength is calculated for the left-out participants in the testing set (sixth box), and the model is applied to generate *d*’ predictions for these left-out subjects.

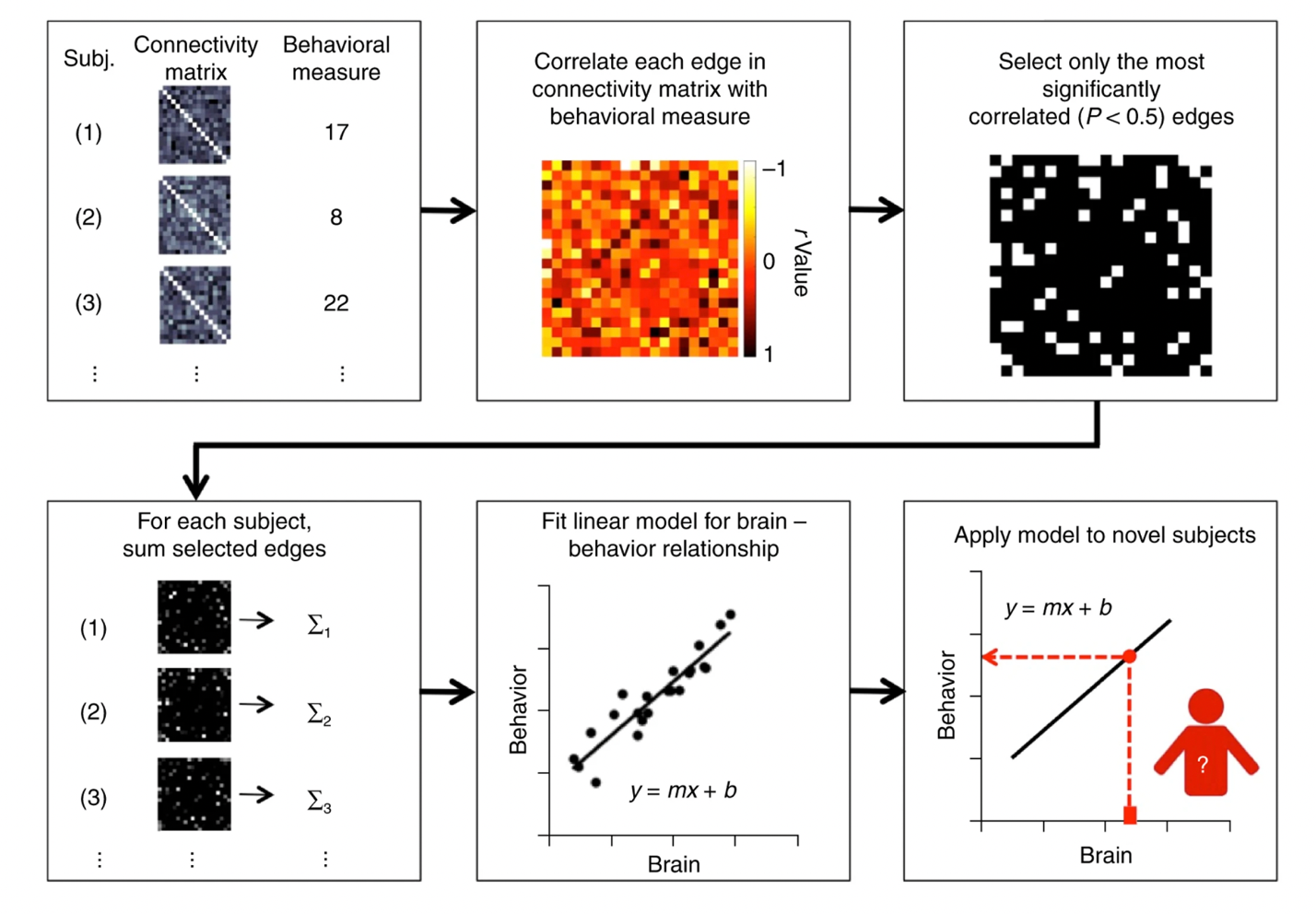

Supplemental Figure 1. A schematic of CPM. Figure reproduced with permission from Shen et al. (2017).

Connectome-based identification (ID)

We performed connectome-based ID (Supplemental Figure 2) as follows. After selecting only the edges in the high and low networks (i.e. the same consensus edges used in the cross-data test), a database is created consisting of all subjects’ matrices from scan 1. In an iterative process, a connectivity matrix from a given subject is then selected from scan 2 and denoted as the target. Pearson correlation coefficients are calculated between the target connectivity matrix and all the matrices in the database. If the highest Pearson correlation coefficient is between the target subject in one session and the same subject in the second session, this is recorded as a correct identification. The process is repeated until identifications have been performed for all subjects and database-target combinations (i.e., each subject serves as a target once in both scan sessions). We average both database-target pairs (because these can be reversed) for a dataset to achieve an average ID rate. We perform this once per sample for the high attention network and once per sample for the low attention network.

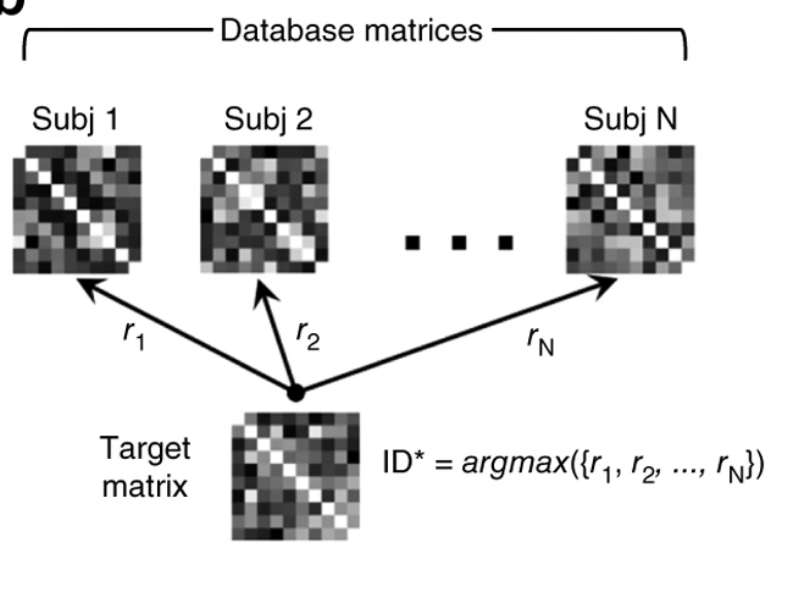

Supplemental Figure 2. A schematic of connectome-based identification. Figure reproduced with permission from Finn et al. (2015).

**Supplemental Results**

External validation of the attention network—results at different thresholds

| **Positive network size** | **Negative network size** | **Combined network size** | **Rho** | ***P*-value** |
| --- | --- | --- | --- | --- |
| 1749 | 1697 | 3446 | 0.66 | 0.0005 |
| 1360 | 1322 | 2682 | 0.62 | 0.001 |
| 1122 | 1083 | 2205 | 0.62 | 0.001 |
| 922 | 896 | 1818 | 0.65 | 0.0006 |
| 744 | 726 | 1470 | 0.65 | 0.0007 |
| 574 | 544 | 1118 | 0.58 | 0.003 |
| 361 | 348 | 709 | 0.50 | 0.01 |
| 34 | 39 | 73 | 0.23 | 0.27 |

Supplemental Table 1. Testing different attention network sizes and determining the relationship to attention score. To generate each summary network, we required that an edge appear in *n*/10 k-folds and *j*/1000 CPM iterations. The variables *n* and *j* were initially set at 3 and 300, respectively. Subsequently, the variables were altered in increments of 1 and 100, respectively. The rows of the table are arranged such that the top row is the most liberal network summarization approach (e.g. an edge mush appear in 3/10 folds and 300/1000 iterations; the bottom of the table is the most stringent case (e.g. 10/10 folds and 1000/1000 iterations). Rho = Spearman’s rho of the correlation between the predicted and observed *d*’ score. Note values in the Rho column are shown with two significant figures.

Neuroanatomy of CPM predictive networks—visualized without correcting for network size

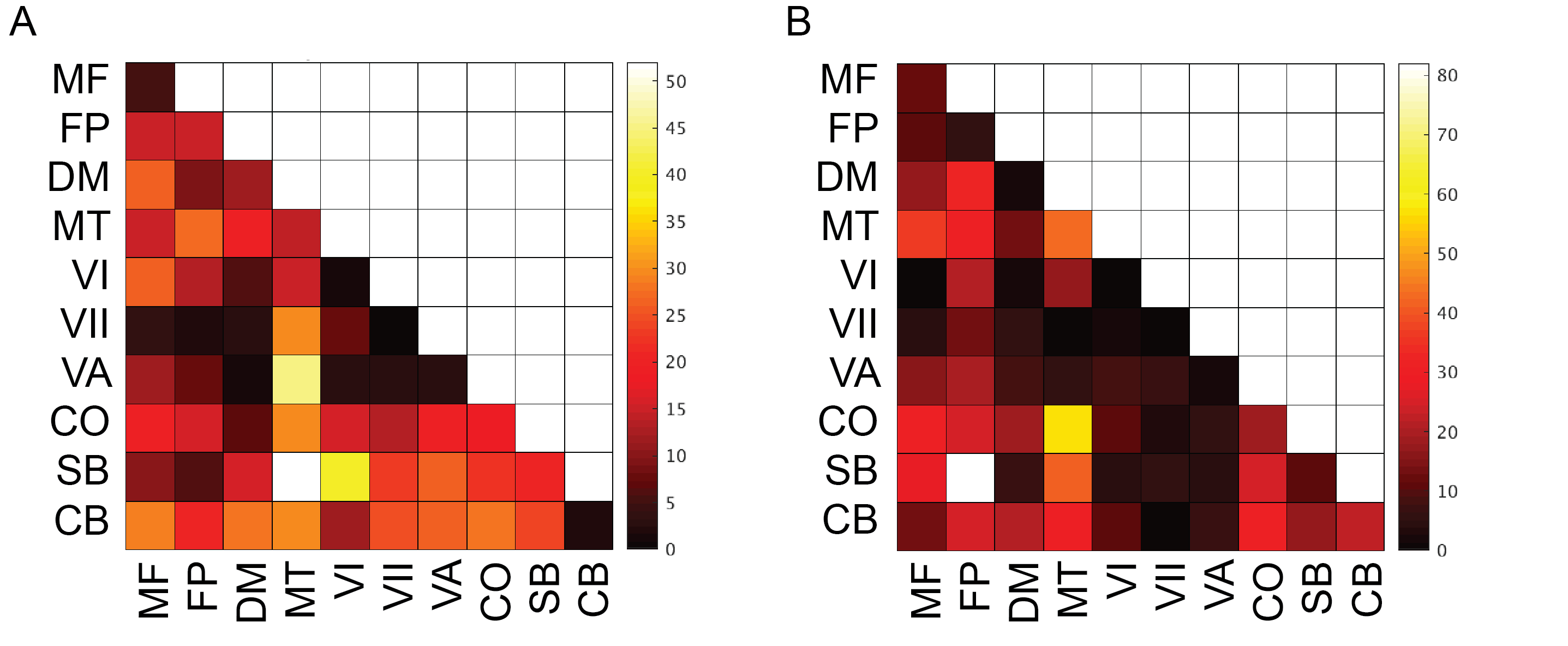

Supplemental Figure 3. Visualizing CPM predictive networks without correcting for network size. A) Results for the high network. B) Results for the low network. Note the color bars of A) and B) are not the same. MF, medial frontal; FP, frontoparietal; DM, default mode; MT, motor; VI, visual I; VII, visual II; VA, visual association; CO, cingulo-opercular; SB, subcortical; CB, cerebellum.

Neuroanatomy of CPM predictive networks—visualized as circle plots without a degree threshold

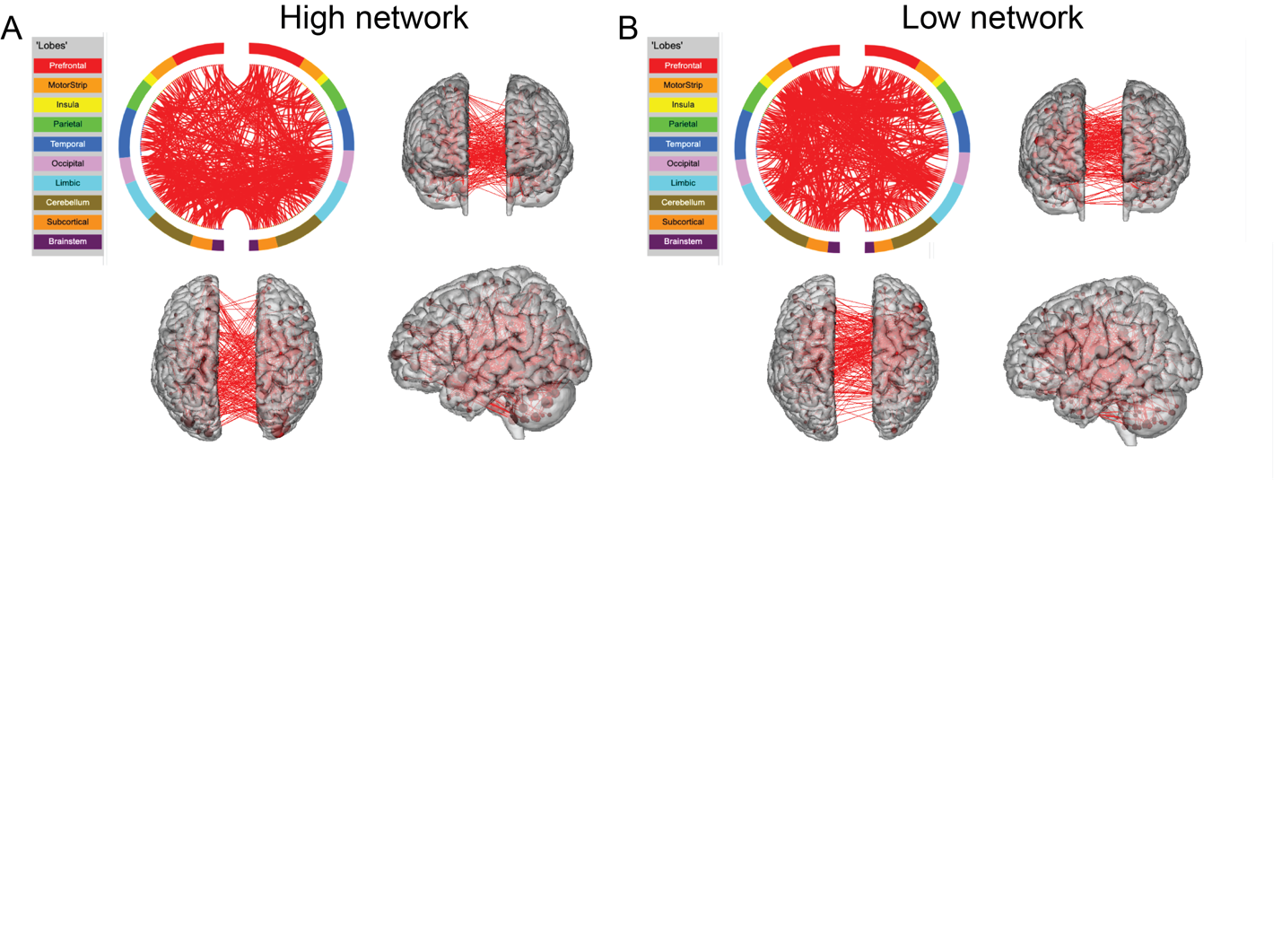

Supplemental Figure 4. Neuroanatomy of CPM predictive networks. A) The consensus high network. B) The consensus low network. For both A) and B): a circle plot is shown to the upper left. The top of the circle represents anterior; the bottom, posterior. The left half of the circle plot corresponds to the left hemisphere of the brain. A legend indicating the approximate anatomic ‘lobe’ is shown to the left. The same edges are plotted in the glass brains as lines connecting different nodes (red circles); in these visualizations, nodes are sized according to degree, the number of edges connected to that node. Note that here we are replotting the data shown in Figure 3A-B, except we have not applied a degree threshold.

In-scanner head motion does not lead to higher within-participant self-correlation scores in the attention network

| **Dataset** | **Correlation coefficient** | ***P*-value** |
| --- | --- | --- |
| UM, high attention network | -0.0471 | 0.8154 |
| UM, low attention network | -0.0451 | 0.8231 |
| Utah, high attention network | -0.051 | 0.8512 |
| Utah, low attention network | 0.1236 | 0.6484 |
| Pitt, high attention network | -0.3636 | 0.0153 |
| Pitt, low attention network | -0.0964 | 0.5337 |

Supplemental Table 2. Pearson correlation coefficient between within-subject self-correlation scores in the attention network and mean in-scanner head motion. Note the only significant correlation (Pitt, high attention network: rho = -0.3636, *P* = 0.0153) is negative, indicating that higher head motion is associated with lower within-subject self-correlation scores (and hence decreases ID rates, in line with previous findings that high motion functional data results in statistically significant ID rates; Horien et al., 2018)

| **Sample, network** | **ID rate of attention network** | **Mean ID rate using edges outside attention network** | ***P*-value** |
| --- | --- | --- | --- |
| UM, high attention network | 0.926 | 0.928 | 0.589 |
| UM, low attention network | 0.815 | 0.927 | 0.999 |
| Utah, high attention network | 0.844 | 0.839 | 0.626 |
| Utah, low attention network | 0.813 | 0.834 | 0.799 |
| Pitt, high attention network | 0.534 | 0.624 | 0.984 |
| Pitt, low attention network | 0.625 | 0.620 | 0.497 |

Supplemental Table 3. The attention network does not differ statistically from other connections in terms of stability. The leftmost column indicates the sample and network used to obtain ID rates. The middle left column indicates the ID rates obtained using the actual attention networks (originally plotted in Figure 4). The middle right column indicates the mean ID rate obtained across 1000 iterations of randomly selected edges outside the attention network. The rightmost column indicates the non-parametric *P*-value obtained by computing the number of times random ID rates were numerically greater than or equal to the actual ID rate obtained using the attention network.
